# Association of *Helicobacter Pylori* Infection and Dyslipidemia Among Individuals Attending General Health Checkup of Tribhuvan University Teaching Hospital

**DOI:** 10.64898/2026.09.16.26363205

**Authors:** Tirtha Man Shrestha, Bishal Budha, Shreeram Paudel, Gaurav Nepal, Subash Wagle, Shrawan Dhungana, Sagar Khadka, Helen Shrestha

**Author notes:** Correspondence, Dr. Bishal Budha, Maharajgunj Medical Campus, Institute of Medicine, Tribhuvan University, Maharajgunj, 44600, Nepal.

## Abstract

**Background:** *Helicobacter pylori* is a highly prevalent chronic infection that induces persistent gastric inflammation and has been increasingly implicated in alterations of lipid metabolism. Several studies suggest an association between *H. pylori* infection and dyslipidemia, potentially contributing to atherosclerosis and cardiovascular disease; however, findings remain inconsistent and causal relationships are debated.

**Objective:** The objective of the study is to determine the association between Helicobacter pylori infection and dyslipidemia.

**Materials and methods:** This study is a cross-sectional study conducted at Tribhuvan University Teaching Hospital (TUTH), Kathmandu, Nepal, a tertiary care referral center. It involved a retrospective analysis of records collected at General Health Checkup (GHC) clinic by a physician. Data collection was done retrospectively from pre-existing medical records. The study population included patients with documented *H. pylori* serological status and available lipid profile results during the study period. Information on demographic characteristics, clinical presentation, medication use, lifestyle factors and known comorbidities was collected using a pretested, standardized questionnaire adapted from literature.

Standardized methods were used for anthropometric measurements. Body mass index (BMI) was calculated as weight (kg) divided by height squared (m^2^).

**Results:** This study included a total of 242 participants and categorized them into three groups based on Helicobacter pylori serology - seronegative, equivocal, and seropositive. In the study, BMI and body weight showed significant positive correlations with LDL (r = 0.208, p = 0.008) and total cholesterol (r = 0.187, p = 0.017). Body weight was positively correlated with total cholesterol (r = 0.204, p = 0.009). No significant correlations were observed between fasting blood glucose and lipid parameters, suggesting that *H. pylori* infection may be associated with increased body weight, though not with overt hyperglycemia.

The current study’s findings showed that there was no statistically significant difference in the mean Fasting Blood Sugar (FBS) levels of those with and without H. pylori infection. (FBG: 5.37 ± 1.20 mmol/L in seronegative participants, 5.29 ± 0.93 mmol/L in equivocal participants, and 5.56 ± 0.92 mmol/L in seropositive participants (*p* = 0.347)).

Among other risk factors, a significant association was found between male sex and *H. pylori* infection (*H. pylori*–seropositive men (49.1%) compared to seronegative males (26.5%), p = 0.004). Smoking status did not substantially differ among patients who tested positive for *H. Pylori*, and alcohol consumption showed a weak correlation (P=0.064).

**Conclusion:** The findings of the study suggest that H. pylori seropositivity is significantly associated with higher body weight and dyslipidemia, particularly elevated TC and LDL cholesterol levels, highlighting the importance of lipid screening and weight reduction in *H. pylori*-positive patients.

## Introduction

One of the most prevalent chronic bacterial diseases in the world, *Helicobacter pylori* (*H. pylori*) is a spiral-shaped, gram-negative, micro-aerophilic bacterium that typically colonizes the stomach mucosa of over half of the population (1–11). Poor sanitation, overcrowding, and a lack of healthcare resources frequently contribute to the oral or fecal routes of H. pylori transmission (8,9,12).

East Asian nations including China, South Korea, and Japan have especially high *H. pylori* prevalences (1). The bacterium can live in the stomach for decades without producing obvious clinical signs, and many people are asymptomatic despite its extensive presence (12). In addition to gastrointestinal problems like gastritis, gastric ulcer and eventually peptic ulcer (5,7–10,12–16), *H. Pylori* can cause extra-gastric symptoms including atherosclerosis, peripheral vascular disease and chronic inflammatory reactions which is a very controversial and hot topic of debate (1,2,4–10,12–14,16).

Mandell et al. originally proposed the link between *H. pylori* and the prevalence of cardiovascular illnesses for the first time in 1994 (9). Since then, a number of investigations, including some more recent ones, have demonstrated that *H. pylori* infection is linked to abnormalities in glucose and lipids, which alters total cholesterol, triglycerides, low-density lipoprotein cholesterol (LDL-c), and high-density lipoprotein cholesterol (HDL-c) and has an impact on the cardiovascular system (2,4,8,17). High levels of total cholesterol (TC), triglycerides (TG), LDL-C, low HDL-C, and elevated body mass index (BMI) have been shown to be substantially linked to coronary heart disease (CHD), which is caused by lipids and lipoproteins and includes CVD, stroke, and atherosclerosis (8,10).

Atherosclerosis is facilitated by dyslipidemia, which is characterized by an increase in any of the blood lipid profiles, including plasma TC, TGs, low-density lipoprotein (LDL-c), or low high-density lipoprotein cholesterol (HDL-c) (2,12,18). Both fundamental (genetic) and secondary causes can contribute to dyslipidemia (2). One of the main causes and risk factors for cardiovascular disease (CVD), which is a leading cause of death worldwide, is atherosclerosis (16). Age, smoking, diabetes, dyslipidemia, hypertension, and chronic inflammation are a few of the particular risk factors (16). Since they can only explain about half of the incidence of atherosclerosis, research into other risk factors for incidence is highly desirable (7).

Due to molecular mimicry between the bacterial structure and the host tissue, *H. pylori* causes chronic inflammation in the stomach. This leads to autoimmune gastritis and issues with lipid metabolism, which in turn causes chronic low-grade activation of the coagulation cascade, an increase in lipid accumulation, and a worsening of atherosclerosis (1,2,6,8,13,18). According to studies, gastrointestinal inflammation can decrease lipid absorption, and lipopolysaccharide’s effect on circulating macrophages causes them to release more free radicals, which oxidize LDL-c and turn macrophages into foam cells two processes thought to be involved in the pathophysiology of atherosclerosis (2,8,18).

The disruption of lipid and lipoprotein metabolism caused by an *H. pylori* infection can lead to decreased levels of apolipoprotein A and high-density lipoprotein cholesterol (HDL-c) and increased levels of triglycerides, total cholesterol, low-density lipoprotein cholesterol (LDL-c), and apolipoprotein B (apo B) in the blood (2,8,9,13,17,18).

Furthermore, despite the very high occurrence of *H. pylori* infection, screening for this infection is not common, and many affected people are not aware that they have it. Determining whether routine screening, early detection, and treatment of *H. pylori* is one of the prevention and therapy options for dyslipidemia and CHD will be made easier by investigating the causal link between the two (4,12). Additionally, this study will give the scientific community evidence-based information about the lipid profiles of H. pylori-infected patients as well as supporting data for policymakers.

## Materials and Methods

### Study design, period and study area

A hospital-based cross-sectional study was conducted at Tribhuvan University Teaching Hospital (TUTH), Kathmandu, Nepal. TUTH is the largest medical college and a major tertiary care referral center in the country. Data were collected respectively from medical record of eligible patients at the General Health Checkup (GHC) clinic from October 1 to December 3o, 2019. The hospital provides comprehensive, modern, and affordable healthcare services while also serving as a leading institution for undergraduate and postgraduate medical education.

Ethical clearance was obtained from the Institutional Review Committee (IRC), Institute of Medicine, Tribhuvan University Teaching Hospital (IOM-TUTH) (Ref. No. 56(6-11) E2 76/77) prior to commencement of the study. As this was a retrospective record-based study, data were obtained from pre-existing medical records, with no direct patient contact or intervention.

### Study Population and inclusive criteria

Patients presenting with chief complaints of dyspepsia, regurgitation, abdominal discomfort, and abdominal fullness for weeks to months were evaluated at the General Health Checkup (GHC) clinic by a physician. The GHC clinic maintains comprehensive medical records of all patients, enabling systematic, accurate, and reliable data collection. As this was a retrospective record-based analysis, there was no direct patient contact or intervention involved.

Data were collected retrospectively from pre-existing medical records. An observational, retrospective, cross-sectional design was adopted, conducted over a defined study period. Random sampling was employed to select eligible patient records. The study population comprised patients with documented *Helicobacter pylori* status and available lipid profile results during the study period.

### Inclusion and Exclusion Criteria

The inclusion criteria consisted of patients with documented positive *Helicobacter pylori* status in the GHC clinic records.

The exclusion criteria were:

1. Incomplete or missing data regarding either *H. pylori* status or lipid profile parameters.
2. Patients already receiving *H. pylori* eradication therapy (triple therapy) at the time of evaluation.

The variables analyzed included demographic characteristics (age and sex), anthropometric measurements (body mass index [BMI]), *Helicobacter pylori* status, lipid profile parameters, and associated comorbidities such as diabetes mellitus (DM), hypertension (HTN), and chronic kidney disease (CKD). Demographic, clinical, and laboratory data were extracted from GHC clinic records using a standardized data collection format. The anticipated duration for data extraction, compilation, and analysis was six months.

### Data collection and measurement

During the study period, data were collected through systematic review of General Health Checkup (GHC) clinic records. A pretested, standardized questionnaire adapted from relevant literature was used to obtain information on demographic characteristics, clinical symptoms (chronic or recurrent upper abdominal pain, dyspepsia, gastroesophageal reflux, and anorexia), medication history (including corticosteroids and oral contraceptive pills), lifestyle factors (alcohol intake, smoking status, and physical activity level), and known comorbid conditions. Data collection was performed by trained personnel to ensure consistency and reliability.

Anthropometric measurements were obtained using standardized techniques. Height was measured barefoot in the erect position, and body weight was recorded using a calibrated weighing scale after removal of heavy clothing and accessories. Body mass index (BMI) was calculated as weight (kg) divided by height squared (m^2^).

Venous blood samples were collected under aseptic conditions for laboratory analysis. *Helicobacter pylori* infection status was determined using serological antibody testing. Biochemical parameters, including fasting lipid profile and blood glucose levels, were retrieved from laboratory records. Lipid profile parameters included triglycerides (TG), low-density lipoprotein cholesterol (LDL-C), high-density lipoprotein cholesterol (HDL-C), and total cholesterol (TC).

Dyslipidemia was defined according to standard clinical criteria: TG >150 mg/dL, LDL-C >130 mg/dL, HDL-C <40 mg/dL, and TC >200 mg/dL.

### Data Quality Assurance

Following data extraction from General Health Checkup (GHC) clinic records, all collected information was independently cross-verified by data collectors and principal investigators to ensure accuracy and completeness. Adherence to the predefined inclusion and exclusion criteria was rigorously maintained through systematic cross-checking of patient records. The structured and comprehensive nature of the GHC clinic documentation system, integral to hospital management and disease surveillance, further contributed to the reliability and quality of the data.

All procedures related to venous blood sample collection, handling, transportation, and laboratory analysis were performed in strict accordance with the standard operating protocols of Tribhuvan University Teaching Hospital (TUTH), thereby ensuring consistency, safety, and analytical validity.

## Results

### Sociodemographic and Behavioral Factors (sheet 1)

A total of 242 participants were enrolled and categorized into three groups based on Helicobacter pylori serology - seronegative, equivocal, and seropositive. Among H. pylori seropositive subjects, males constituted 49.1%, compared to 26.5% in the seronegative group - a statistically significant difference (p = 0.004). Alcohol consumption was more common among *H. pylori* seropositive individuals compared with seronegative ones, though the difference was not statistically significant (p = 0.064). Smoking, HTN, DM, OCP consumption and menopause was not significantly associated with *H. pylori* infection.

| Variable | Category | H. pylori seronegative | Equivocal | H. pylori seropositive | P value |
| --- | --- | --- | --- | --- | --- |
| **SEX** |  |  |  |  | **0.004** |
|  | F | 50 (73.5%) | 8 (72.7%) | 83 (50.9%) |  |
|  | M | 18 (26.5%) | 3 (27.3%) | 80 (49.1%) |  |
| **ALCOHOL** |  |  |  |  | **0.064** |
|  | No | 57 (83.8%) | 7 (63.6%) | 115<br>(70.6%) |  |
|  | Yes | 11 (16.2%) | 4 (36.4%) | 48 (29.4%) |  |
| **SMOKING** |  |  |  |  | **0.829** |
|  | No | 53 (77.9%) | 8 (72.7%) | 122<br>(74.8%) |  |
|  | Yes | 15 (22.1%) | 3 (27.3%) | 41 (25.2%) |  |
| **HTN** |  |  |  |  | **0.423** |
|  | No | 52 (76.5%) | 10<br>(90.9%) | 114<br>(69.9%) |  |
|  | Y | 0 (0.0%) | 0 (0.0%) | 1 (0.6%) |  |
|  | Yes | 16 (23.5%) | 1 (9.1%) | 48 (29.4%) |  |
| **DM** |  |  |  |  | **0.114** |
|  | No | 65 (95.6%) | 10<br>(90.9%) | 142<br>(87.1%) |  |
|  | Yes | 3 (4.4%) | 1 (9.1%) | 21 (12.9%) |  |
| **CKD** |  |  |  |  |  |
|  | No | 68 (100.0%) | 11<br>(100.0%) | 163<br>(100.0%) |  |
| **CLD** |  |  |  |  |  |
|  | No | 68 (100.0%) | 11<br>(100.0%) | 163<br>(100.0%) |  |
| **MALIGNANCY** |  |  |  |  |  |
|  | No | 68 (100.0%) | 11<br>(100.0%) | 163<br>(100.0%) |  |
| **OCP** |  |  |  |  | **0.674** |
|  | No | 56 (82.4%) | 8 (72.7%) | 131<br>(80.4%) |  |
|  | Yes | 12 (17.6%) | 3 (27.3%) | 32 (19.6%) |  |
| **MENOPAUSE** |  |  |  |  | **0.057** |
|  | No | 49 (72.1%) | 8 (72.7%) | 138<br>(84.7%) |  |
|  | Yes | 19 (27.9%) | 3 (27.3%) | 25 (15.3%) |  |
| **FAMILY DYSLIPIDEMIA** |  |  |  |  | **0.049** |
|  | No | 65 (95.6%) | 8 (72.7%) | 149 (91.4%) |  |
|  | Yes | 3 (4.4%) | 3 (27.3%) | 14 (8.6%) |  |

### Baseline Characteristics (Sheet 2)

There was no significant difference in the participants’ mean age across the groups (p = 0.865*). H. pylori* seropositive individuals had a substantially greater mean body weight (68.55 ± 8.55 kg) than seronegative persons (65.07 ± 9.10 kg; p = 0.013). Height, BMI, and fasting blood glucose levels did not differ significantly across the groups (p > 0.05). This suggests that *H. pylori* infection may be associated with increased body weight, though not necessarily with overt hyperglycemia.

Participants who tested positive for *H. pylori* had noticeably greater body weights (Sheet 2). The association between *H. pylori* infection and dyslipidemia may be mediated through its association with increased body weight or BMI, as body weight and BMI positively correlate with LDL and TC (Sheet 4). Although the illness may not directly raise lipid levels, its presence in high BMI people may indicate metabolic alterations that increase the risk of elevated LDL and TC.

| Variable | seronegative | equivocal | seropositive | P_value |
| --- | --- | --- | --- | --- |
| AGE | 45.31 ± 14.47 | 43.09 ± 9.08 | 44.66 ± 13.28 | 0.865 |
| WEIGHT | 65.07 ± 9.10 | 70.36 ± 8.27 | 68.55 ± 8.55 | 0.013 |
| HEIGHT<br>m | 1.58 ± 0.08 | 1.59 ± 0.07 | 1.58 ± 0.18 | 0.930 |
| BMI | 26.00 ± 3.50 | 27.90 ± 2.48 | 65.01 ± 342.12 | 0.605 |
| FBG | 5.37 ± 1.20 | 5.29 ± 0.93 | 5.56 ± 0.92 | 0.347 |
| TG | 1.92 ± 1.07 | 1.85 ± 0.80 | 2.07 ± 1.07 | 0.522 |
| LDL | 2.61 ± 0.88 | 3.46 ± 1.15 | 2.94 ± 0.97 | 0.006 |
| HDL | 1.27 ± 0.70 | 1.36 ± 0.31 | 1.24 ± 0.53 | 0.765 |
| TC | 4.75 ± 0.98 | 5.25 ± 0.88 | 5.14 ± 1.00 | 0.022 |

### Association Between Dyslipidemia and Risk Factors (sheet 3)

54.5% of men and 43.3% of women had hypercholesterolemia (TC > 200 mg/dl). Alcohol use was substantially linked to hypercholesterolemia in crude analysis (cOR = 2.15; 95% CI: 1.20–3.92; p = 0.011). After controlling for covariates, this relationship was not significant (aOR = 1.00; 95% CI: 0.37–2.66; p = 0.996). Sex and other factors did not show any significant adjusted relationships.

| Variable | Category | <200<br>mg/dl<br>n (%) | ≥200<br>mg/dl<br>n (%) | cO<br>R | cO<br>R<br>95<br>%<br>CI | cOR<br>p | aO<br>R | aO<br>R<br>95<br>%<br>CI | aOR<br>p |
| --- | --- | --- | --- | --- | --- | --- | --- | --- | --- |
| SEX | M | 46<br>(45.5<br>%) | 55<br>(54.5<br>%) | 1.0<br>0 | Ref | - | 1.0<br>0 | Ref | - |
|  | F | 80<br>(56.7<br>%) | 61<br>(43.3<br>%) | 0.6<br>4 | 0.38<br>–<br>1.06 | 0.086 | 0.9<br>5 | 0.43<br>–<br>2.10 | 0.901 |
| ALCOHOL | No | 102<br>(57%) | 77<br>(43%) | 1.0<br>0 | Ref | - | 1.0<br>0 | Ref | - |
|  | Yes | 24<br>(38.1<br>%) | 39<br>(61.9<br>%) | 2.1<br>5 | 1.20<br>–<br>3.92 | **0.011** | 1.0<br>0 | 0.37<br>–<br>2.66 | 0.996 |
| SMOKING | No | 105<br>(57.4<br>%) | 78<br>(42.6<br>%) | 1.0<br>0 | Ref | - | 1.0<br>0 | Ref | - |
|  | Yes | 21<br>(35.6<br>%) | 38<br>(64.4<br>%) | 2.4<br>4 | 1.34<br>–<br>4.54 | **0.004** | 2.0<br>3 | 0.76<br>–<br>5.63 | 0.163 |
| HTN | No | 98<br>(55.7<br>%) | 78<br>(44.3<br>%) | 1.0<br>0 | Ref | - | 1.0<br>0 | Ref | - |
|  | Yes | 28<br>(42.4<br>%) | 38<br>(57.6<br>%) | 1.7<br>1 | 0.97<br>–<br>3.04 | 0.067 | 1.1<br>9 | 0.57<br>–<br>2.48 | 0.644 |
| DM | No | 113<br>(52.1<br>%) | 104<br>(47.9<br>%) | 1.0<br>0 | Ref | - | 1.0<br>0 | Ref | - |
|  | Yes | 13<br>(52%) | 12<br>(48%) | 1.0<br>0 | 0.43<br>–<br>2.31 | 0.994 | 0.8<br>5 | 0.31<br>–<br>2.32 | 0.754 |
| OCP | No | 94<br>(48.2<br>%) | 101<br>(51.8<br>%) | 1.0<br>0 | Ref | - | 1.0<br>0 | Ref | - |
|  | Yes | 32<br>(68.1<br>%) | 15<br>(31.9<br>%) | 0.4<br>4 | 0.22<br>–<br>0.84 | **0.016** | 0.3<br>8 | 0.14<br>–<br>1.00 | **0.049** |
| MENOPAUSE | No | 105<br>(53.8<br>%) | 90<br>(46.2<br>%) | 1.0<br>0 | Ref | - | 1.0<br>0 | Ref | - |
|  | Yes | 21<br>(44.7<br>%) | 26<br>(55.3<br>%) | 1.4<br>4 | 0.76<br>–<br>2.76 | 0.260 | 2.4<br>1 | 0.81<br>–<br>7.21 | 0.113 |
| FAMILY_DYSLIPID<br>EMIA | No | 115<br>(51.8<br>%) | 107<br>(48.2<br>%) | 1.0<br>0 | Ref | - | 1.0<br>0 | Ref | - |
|  | Yes | 11<br>(55%) | 9<br>(45%) | 0.8<br>8 | 0.34<br>–<br>2.21 | 0.784 | 0.6<br>7 | 0.21<br>–<br>2.02 | 0.474 |
| HPYLORI_CAT | seronegati<br>ve | 47<br>(69.1<br>%) | 21<br>(30.9<br>%) | 1.0<br>0 | Ref | - | 1.0<br>0 | Ref | - |
|  | equivocal | 7<br>(63.6<br>%) | 4<br>(36.4<br>%) | 1.2<br>8 | 0.31<br>–<br>4.72 | 0.717 | 1.6<br>7 | 0.32<br>–<br>8.27 | 0.527 |
|  | seropositi<br>ve | 72<br>(44.2<br>%) | 91<br>(55.8<br>%) | 2.8<br>3 | 1.57<br>–<br>5.23 | **<0.0**<br>**01** | 3.6<br>2 | 1.85<br>–<br>7.36 | **<0.0**<br>**01** |
| AgeCategory | <40 | 46<br>(52.3<br>%) | 42<br>(47.7<br>%) | 1.0<br>0 | Ref | - | 1.0<br>0 | Ref | - |
|  | >50 | 33<br>(47.8<br>%) | 36<br>(52.2<br>%) | 1.1<br>9 | 0.64<br>–<br>2.25 | 0.580 | 0.5<br>6 | 0.22<br>–<br>1.41 | 0.224 |
|  | 41–45 | 17<br>(50%) | 17<br>(50%) | 1.1<br>0 | 0.49<br>–<br>2.43 | 0.822 | 1.5<br>4 | 0.59<br>–<br>4.07 | 0.381 |
|  | 46–50 | 22<br>(59.5<br>%) | 15<br>(40.5<br>%) | 0.7<br>5 | 0.34<br>–<br>1.62 | 0.462 | 0.5<br>4 | 0.22<br>–<br>1.30 | 0.175 |
| BMI_Category | Normal | 34<br>(49.3<br>%) | 35<br>(50.7<br>%) | 1.0<br>0 | Ref | - | 1.0<br>0 | Ref | - |
|  | Obese | 18<br>(41.9<br>%) | 25<br>(58.1<br>%) | 1.3<br>5 | 0.63<br>–<br>2.93 | 0.445 | 1.1<br>9 | 0.48<br>–<br>2.97 | 0.701 |
|  | Overweig<br>ht | 72<br>(56.2<br>%) | 56<br>(43.8<br>%) | 0.7<br>6 | 0.42<br>–<br>1.36 | 0.349 | 0.7<br>0 | 0.35<br>–<br>1.39 | 0.310 |
|  | Underwei<br>ght | 2<br>(100<br>%) | 0<br>(0%) | 0.0<br>0 | 0.31<br>–<br>4.72 | 0.981 | 0.0<br>0 | 0 | 0.988 |

### Correlation Between Anthropometric/Biochemical Predictors and Lipid Profile (Sheet 4)

BMI and body weight showed significant positive correlations with LDL (r = 0.208, p = 0.008) and total cholesterol (r = 0.187, p = 0.017). Similarly, weight correlated positively with total cholesterol (r = 0.204, p = 0.009). Height showed a weak but statistically significant positive correlation with triglyceride levels (r = 0.173, p = 0.028). No significant correlations were observed between fasting blood glucose and lipid parameters. Age, and height show no significant correlation with either LDL or TC. This suggests that *H. pylori* infection may be associated with increased body weight, though not necessarily with overt hyperglycemia.

| Predictor | TG | LDL | HDL | TC |
| --- | --- | --- | --- | --- |
| AGE | -0.060<br>(p=0.444) | -0.060<br>(p=0.446) | 0.028<br>(p=0.720) | -0.116<br>(p=0.140) |
| BMI | -0.136<br>(p=0.084) | 0.208<br>(p=0.008) | -0.139<br>(p=0.077) | 0.187<br>(p=0.017) |
| FBG | 0.032<br>(p=0.687) | -0.078<br>(p=0.325) | -0.130<br>(p=0.098) | -0.122<br>(p=0.123) |
| WEIGHT | 0.051<br>(p=0.517) | 0.159<br>(p=0.043) | -0.138<br>(p=0.079) | 0.204<br>(p=0.009) |
| HEIGHT<br>m | 0.173<br>(p=0.028) | -0.082<br>(p=0.297) | -0.026<br>(p=0.740) | -0.022<br>(p=0.786) |
| SEX | 0.172<br>(p=0.028) | -0.002<br>(p=0.976) | 0.017<br>(p=0.833) | 0.117<br>(p=0.138) |
| ALCOHOL | 0.100<br>(p=0.206) | 0.089<br>(p=0.259) | -0.001<br>(p=0.992) | 0.134<br>(p=0.090) |
| SMOKING | 0.155<br>(p=0.048) | 0.169<br>(p=0.031) | 0.026<br>(p=0.743) | 0.232<br>(p=0.003) |
| HTN | -0.057<br>(p=0.471) | -0.011<br>(p=0.885) | 0.038<br>(p=0.626) | -0.019<br>(p=0.815) |
| DM | -0.035<br>(p=0.659) | -0.090<br>(p=0.254) | 0.063<br>(p=0.424) | -0.019<br>(p=0.810) |
| CKD |  |  |  |  |
| CLD |  |  |  |  |
| MALIGNANCY |  |  |  |  |
| OCP | -0.006<br>(p=0.936) | -0.065<br>(p=0.408) | 0.045<br>(p=0.565) | -0.121<br>(p=0.125) |
| MENOPAUSE | -0.156<br>(p=0.046) | 0.057<br>(p=0.473) | 0.060<br>(p=0.450) | 0.030<br>(p=0.700) |
| FAMILY<br>DYSLIPIDEMIA | 0.006<br>(p=0.943) | -0.151<br>(p=0.055) | 0.192<br>(p=0.014) | -0.050<br>(p=0.525) |

## Discussion

Over 80% of people in underdeveloped nations suffer from *H. pylori* infection, one of the most common bacterial illnesses worldwide (19). The study population in our study had an overall seropositive prevalence of 67.4%.

Dyslipidemia is a set of plasma lipid and lipoprotein abnormalities that are metabolically linked. It is characterized by low HDL-c and elevated LDL-c, TGs, and total cholesterol (TC) levels. High levels of TC, TG, LDL-c, low HDL-c, and an elevated body mass index (BMI) have been demonstrated to be strongly linked with Coronary Heart Disease (CHD) (20)

There is growing evidence in the literature that showed link between *H. pylori* and extra-gastric diseases. *H. pylori* infection has been proven to affect the cardiovascular system, resulting in alterations in total cholesterol, triglycerides, LDL-c, and HDL cholesterol (19,21,22). Patients who have tested positive for *H. pylori* are more likely to develop dyslipidemia and may have altered blood lipid profile than seronegative patient(20).

Our current study revealed that H pylori infection and dyslipidemia are related through increased body weight. Research conducted in different part of world showed that the infected group had higher levels of TG, TC, and LDL but our results suggest that *H. pylori* infection is associated with increase in body weight and associated significantly increase in LDL and TC but increase in TG was not statistically significant (20,23–26). Our findings corroborate a study carried out in Ethiopia to assess the lipid profile of H pylori patients, which discovered that *H. pylori* infection was linked to an increase in blood cholesterol, TG, and LDL-C but a decrease in serum HDL-C that was not statistically significant.(27) and same for study done in Spain (28) and Japan(29).This may be the result of cytokines, particularly tumor necrosis factor, which increases the production of free radicals and inhibits lipoprotein lipase. This consequently promotes LDL-c oxidation, a crucial step in the development of atherosclerosis.(30–32)

*H. pylori* positive individuals in our study had significantly higher serum TC (P = 0.022) and LDL (P = 0.006) concentrations than *H. pylori* negative individuals, which may raise the risk of CHD. This is consistent with a study done in Finland (33).

The current study’s findings showed that there was no statistically significant difference in the mean Fasting Blood Sugar (FBS) levels of those with and without H. pylori infection. (FBG : 5.37 ± 1.20 mmol/L in seronegative participants, 5.29 ± 0.93 mmol/L in equivocal participants, and 5.56 ± 0.92 mmol/L in seropositive participants (*p* = 0.347)) which contrasted to the study done in Iran (34,35) and study done in Romania (36) which showed that fasting blood sugar is higher in *H. pylori* seropositive patient. Further correlation analysis demonstrated no significant association between fasting blood glucose and lipid parameters, including LDL cholesterol and total cholesterol (FBG vs LDL: r = −0.078, *p* = 0.325; FBG vs TC: r = −0.122, *p* = 0.123). This implies that lipid abnormalities in the presence of H. pylori infection in this population were unaffected by glycemic state.

In this cross-sectional study, we found that H pylori seropositivity was associated with higher mean body weight compared with seronegative participants (68.6 ± 8.6 kg vs 65.1 ± 9.1 kg; p = 0.013). BMI and body weight were positively linked with LDL and TLC, despite the fact that BMI did not significantly differ across groups in our study. Similar research was conducted in China(37), Iran (38) which emphasized that *H. pylori* infection was significantly and positively associated with overweight.

Our results imply that, rather than through direct metabolic effects, *H. pylori* infection may indirectly contribute to an elevated lipid profile by its connection with higher body weight and adiposity, nevertheless, studies conducted in Ethiopia and Korea indicate a direct association (19,39).

Regarding the other risk variables, our study found a significant correlation between male sex and *H. pylori* infection (*H. pylori*–seropositive men (49.1%) compared to seronegative males (26.5%), p = 0.004). Smoking did not substantially differ among patients who tested positive for *H. Pylori*, and alcohol consumption showed a weak correlation (P=0.064).

According to earlier research, the main ways that *H. pylori* causes cardiovascular diseases include the activation of inflammatory mediators, the release of toxins, proinflammatory factors, autoimmune reactions, immune system dysfunction, and changes to lipid and iron metabolism (40,41)In opposite to the facts, the idea that *H. pylori* is linked to changes in lipid profiles was not supported by several investigations. For example, according to a cohort research by *Elizalde et al*., blood lipid levels were unaffected by *H. pylori* infection prior to and three months following eradication therapy with a low treatment rate. (42)

A successful *H. pylori* eradication treatment can lower the risk of high LDL and low HDL, according to some research (43). According to a Hungarian study, some *H. pylori*-infected patients had long-term atherogenic complications, which can increase numerous other complex clinical conditions like peripheral artery occlusive disease, coronary heart disease, and brain stroke. (44)

As this was a retrospective, record based study, strong conclusions cannot be drawn using the available data. The number of H. Pylori positive patients was limited and the sample size was small, so the result cannot be generalized to a larger population. Additionally, not all the the lipid parameters could be included in the study due to unavailability of data, such as lipoprotein a. Also, the causal relationships between the studied parameters cannot be established through this cross-sectional nature of the study.

## Conclusion

In conclusion, this cross-sectional study demonstrates that *Helicobacter pylori* seropositivity is significantly associated with male sex, higher body weight, and adverse lipid profiles, particularly elevated total cholesterol and LDL cholesterol. Dyslipidemia was present in nearly half of seropositive individuals, indicating that *H. pylori* infection may contribute to atherogenic risk primarily through adiposity-mediated pathways rather than direct metabolic or glycemic effects. BMI and body weight correlated positively with LDL and total cholesterol, whereas smoking, alcohol use, and fasting glucose showed no significant associations. These findings underscore the importance of targeted lipid screening and weight management in *H. pylori*-positive patients and highlight the need for prospective studies to clarify causality and evaluate the impact of eradication therapy on lipid profiles and cardiovascular risk.

## Data Availability

Data is available on the request.

## Declaration

## Acknowledgement

We express our sincere thanks to Department of General Practice and Emergency Medicine, Tribhuvan University Teaching Hospital, Kathmandu for giving us the opportunity to conduct this study in the premises. Special acknowledgment to all who were directly or indirectly involved in this research for their valuable help.

## Ethical clearance

Ethical clearance was obtained from IRC IOM TUTH (Ref no: 56(6-11) E2 76/77), before conducting this study and a written consent was also taken from department of General Practice and Emergency Medicine, Tribhuvan University Teaching Hospital before data collection.

## Consent for study

Written informed consent was taken from each participants

## Consent for publication

It was taken from researchers as mentioned in authorship declaration form.

## Author Contributions

TMS: Conceptualization, Writing and Editing Original Draft

BB: Writing and Editing of Original Draft, Revision, Design of Work

SP: Data Collection, Writing and Editing of Original Draft

GN: Conceptualization, Methodology

SW: Data Analysis

SD: Writing and Editing of Original Draft

SK: Writing and Editing of Original Draft

HS: Writing and Editing of Original Draft

